# The psychosocial impact of the COVID-19 pandemic on 4,378 UK healthcare workers and ancillary staff: initial baseline data from a cohort study collected during the first wave of the pandemic

**DOI:** 10.1101/2021.01.21.20240887

**Authors:** Danielle Lamb, Sam Gnanapragasam, Neil Greenberg, Rupa Bhundia, Ewan Carr, Matthew Hotopf, Reza Razavi, Rosalind Raine, Sean Cross, Amy Dewar, Mary Docherty, Sarah Dorrington, Stephani L Hatch, Charlotte Wilson-Jones, Daniel Leightley, Ira Madan, Sally Marlow, Isabel McMullen, Anne Marie Rafferty, Martin Parsons, Catherine Polling, Danai Serfioti, Peter Aitken, Veronica French, Helen Gaunt, Joanna Morris-Bone, Rachel Harris, Chloe Simela, Sharon A M Stevelink, Simon Wessely, On behalf of the NHS CHECK consortium

## Abstract

**Objectives:** This study reports preliminary findings on the prevalence of, and factors associated with, mental health and wellbeing outcomes of healthcare workers during the early months (April-June) of the COVID-19 pandemic in the UK.

**Methods:** Preliminary cross-sectional data were analysed from a cohort study (n=4,378). Clinical and non-clinical staff of three London-based NHS Trusts (UK), including acute and mental health Trusts, took part in an online baseline survey. The primary outcome measure used is the presence of probable common mental disorders (CMDs), measured by the General Health Questionnaire (GHQ-12). Secondary outcomes are probable anxiety (GAD-7), depression (PHQ-9), Post-Traumatic Stress Disorder (PTSD) (PCL-6), suicidal ideation (CIS-R), and alcohol use (AUDIT). Moral injury is measured using the Moray Injury Event Scale (MIES).

**Results:** Analyses showed substantial levels of CMDs (58.9%, 95%CI 58.1 to 60.8), and of PTSD (30.2%, 95%CI 28.1 to 32.5) with lower levels of depression (27.3%, 95%CI 25.3 to 29.4), anxiety (23.2%, 95%CI 21.3 to 25.3), and alcohol misuse (10.5%, 95%CI, 9.2 to 11.9). Women, younger staff, and nurses tended to have poorer outcomes than other staff, except for alcohol misuse. Higher reported exposure to moral injury (distress resulting from violation of one’s moral code) was strongly associated with increased levels of CMDs, anxiety, depression, PTSD symptoms, and alcohol misuse.

**Conclusions:** Our findings suggest that mental health support for healthcare workers should consider those demographics and occupations at highest risk. Rigorous longitudinal data are needed in order to respond to the potential long-term mental health impacts of the pandemic.

**Highlights:** *What is already known about this subject?:*

- Large-scale population studies report increased prevalence of depression, anxiety, and psychological distress during the COVID-19 pandemic.
- Evidence from previous epidemics indicates a high and persistent burden of adverse mental health outcomes among healthcare workers.

*What are the new findings?:*

- Substantial levels of probable common mental disorders and post-traumatic stress disorder were found among healthcare workers.
- Groups at increased risk of adverse mental health outcomes included women, nurses, and younger staff, as well as those who reported higher levels of moral injury.

*How might this impact on policy or clinical practice in the foreseeable future?:*

- The mental health offering to healthcare workers must consider the interplay of demographic, social, and occupational factors.
- Additional longitudinal research that emphasises methodological rigor, namely with use of standardised diagnostic interviews to establish mental health diagnoses, is necessary to better understand the mental health burden, identify those most at risk, and provide appropriate support without pathologizing ordinary distress responses.

## Introduction

The NHS has faced extraordinary pressures during the COVID-19 pandemic. NHS clinical and non-clinical healthcare workers (HCWs) have had to contend with a range of significant stressors including the risk of infection, workload changes, sleep deprivation, loss of colleagues, and sometimes providing care in less than adequate settings, while reporting insufficient access to personal protective equipment (PPE). All of these stressors may contribute to an increase in the prevalence of common mental disorders (CMDs), as well as exposing HCWs to so called ‘moral injury’, the distress resulting from violation of one’s moral code. For example, HCWs may feel betrayed by their co-workers or others outside the healthcare service who they once trusted, provided patient care in such a way that it violated their own moral codes or values, or saw things in the healthcare service that they felt were morally wrong.[1]

In addition to workplace-based stressors, HCWs are also vulnerable to wider socioeconomic stressors such as loss of social support, financial difficulties and infection/death of loved ones. Concerns have been raised about the potential psychosocial toll on this population during COVID-19, based on evidence from previous epidemics, mainly Severe Acute Respiratory Syndrome (SARS), Middle East Respiratory Syndrome (MERS) and Ebola. Symptoms of anxiety, depression and post-traumatic stress disorder (PTSD) have been reported as adverse outcomes for HCWs during, and in the years following, those epidemics.[1–3]

Self-reported mental health has been deteriorating across the UK during the pandemic, with large-scale population studies reporting increased prevalence of depression and anxiety.[4–6] However, there is conflicting evidence about whether the psychosocial impact of the COVID-19 pandemic has been more detrimental for HCWs than for the general population.

A significant limitation of existing research related to the mental health of HCWs during COVID-19 is the use of cross-sectional study designs and convenience samples recruited via social media, with no clear sampling frame or ability to calculate a response rate and assess bias.[7] Our study (NHS CHECK) aims to remedy these limitations using a cohort design with a defined source population including all NHS staff (not just clinical staff). The detailed information we hold about our population is important in allowing us to assess the representativeness of our sample, and understand and mitigate the impact of non-response bias. NHS CHECK is investigating CMDs and moral injury, as well as commonly reported adverse mental health outcomes (e.g. anxiety, depression, PTSD, suicidal ideation, and alcohol misuse). We report on preliminary baseline data collected during the initial lockdown period in the UK (April-June 2020), describing a cohort of HCWs from a known population of all staff in the first three of the participating Trusts. Here we report associations between sociodemographic and occupational factors with adverse mental health outcomes in HCWs.

Our research questions were:

1. What is the sociodemographic, occupational, and mental health profile of our sample of HCWs?
2. What factors are associated with probable CMDs as measured by the GHQ-12?
3. What factors are associated with other adverse mental health outcomes as measured by the GAD-7, PHQ-9, AUDIT, PCL-6, and CIS-R?

## Method

### Study design and participants

The sample for NHS CHECK includes all staff working in participating NHS Trusts, including medical, nursing, midwifery, allied health professionals, support, administrative, management, and students who volunteered or were fast-tracked into clinical roles during the COVID-19 pandemic. In this paper we report the results from the initial lockdown period of the pandemic (April-June 2020) during which there were ongoing high levels of COVID-19 hospital admissions and mortality. At that time, the study had been launched across three NHS Hospital trusts in South East London serving two of the Greater London boroughs that were in the top ten for highest rates of COVID-19 related deaths during this period.[8]

### Procedures

We identified eligible participants via the Trusts’ human resources (HR) systems. We used the Trusts’ existing group emails to distribute communications to NHS staff explaining and promoting the study. A dedicated NHS CHECK recruitment email was sent by senior management explaining the study, with a link to the study website. A variation of this email was sent repeatedly during baseline recruitment. We also used existing team-based local contacts established through in-reach staff support, for example staff support teams/leads, chief nursing officers, medical directors, occupational health departments, trade union representatives, and wellbeing hub users. NHS CHECK was promoted during team briefings, included in Trust newsletters, news items on Trust intranet websites, closed social media groups and screen savers on Trust computers etc.

### Data collection and materials

A survey was developed with input from the NHS CHECK Patient and Public Involvement (PPI) advisory group and we tested the acceptability of the questions, materials, and data collection procedures with this small informal reference group of front-line staff (psychologists, managers, administrators, security staff, intensivists, and trainees) and refined it accordingly. Participation in the baseline survey of NHS CHECK involved completing a short (5-10 minutes) online questionnaire that was hosted using Qualtrics questionnaire software, with the option to complete an additional, longer (10-15 minutes) questionnaire if they wished. Participant information provided on the study website reinforced that participation was voluntary and confidential. Participants provided online informed consent before starting the online survey. NHS CHECK was launched on the 24^th^ of April 2020, 5 weeks after the initial lockdown in the UK began (23/03/20).

#### Short baseline questionnaire

The short baseline questionnaire collected a variety of information, including 1) contact details, 2) occupational information (e.g. occupational group, length of professional registration), and 3) socio- demographic characteristics (e.g. age, sex, ethnicity, marital status). The primary outcome was prevalence of probable CMDs, which were assessed using the 12-item General Health Questionnaire (GHQ-12), with a cut-off score of 4 or more indicating ‘caseness’ (indicating increased probability of experiencing a recognised mental disorder) of a CMD.[9]

#### Longer baseline questionnaire

If participants opted to fill in the longer baseline survey, the following validated measures on mental health were used as secondary outcomes, with binary outcomes variables defined using the following cut-off scores for caseness: the 7-item Generalised Anxiety Disorder (GAD) scale to measure probable moderate anxiety disorder with a cut-off score of 10 or more;[10] the 9-item Patient Health Questionnaire (PHQ-9) to measure probable moderate depression with a cut-off score of 10 or more;[11] the 10-item Alcohol Use Disorder Identification Test to measure alcohol consumption with a cut-off score of 8 or more indicative of hazardous drinking;[12] and the 6-item Post-Traumatic Stress Disorder checklist (PCL-6) civilian version to measure PTSD[13] with a score of 14 or more indicating the presence of probable PTSD.

In addition, the 9-item Moral Injury Event Scale (MIES) to measure moral injury,[14] with a higher score indicating more exposure to morally injurious events (possible score range 9 – 54). For the purpose of analysis, the MIES was split into tertiles (lower: 9; middle: 10-16; upper: 17-54). We also assessed suicidal ideation, using items related to suicidal thoughts, suicide attempts and self-harm derived from the Clinical Interview Schedule (CIS-R).[15] The following items were asked: “Have you ever thought of taking your life, even though you would not actually do it?”; Have you ever made an attempt to take your life, by taking an overdose of tablets or in some other way?”; and “Have you ever deliberately harmed yourself in any way but not with the intention of killing yourself?”. Response options were, “Yes, in the past 2 months”, “Yes, but not in the past 2 months”, or “No”.

### Statistical analysis

Aggregate-level population data were provided by participating Trust HR departments including information on the number of employees and the age, sex, and ethnicity composition of the workforce. From this, we calculated a response rate for each Trust. Response weights were generated using a raking algorithm based on age, sex and ethnicity. For weighting purposes only, missing demographic data were imputed using multiple imputation. Missingness was no more than 6%, 2%, and 2% for age, sex, and ethnicity, respectively, before imputation.

The analysis proceeded in five stages. First, we described the representativeness of the sample by comparing composition in terms of age, sex, and ethnicity with the HR population data using frequencies and percentages. Second, we summarized survey participants with appropriate descriptive statistics for each variable (frequencies and weighted percentages for categorical variables, mean and standard deviation for continuous variables). Third, we described differences between individuals who completed both the ‘short’ and ‘long’ baseline surveys versus those completing only the ‘short’ survey. Fourth, we summarized the weighted prevalence of the primary (GHQ) and secondary outcomes (GAD-7, PHQ-9, AUDIT, PCL-6, MIES, and CIS-R) for all participants and stratified by socio-demographic and occupational factors. Finally, we used multivariable binary logistic regression models to explore relationships of socio-demographic and occupational factors (age, sex, ethnicity, role), as well as moral injury with the primary and secondary outcomes, adjusting for all covariates in the models. All analyses were conducted using Stata 16.

### Ethical approval

Ethical approval for the study was granted by the Health Research Authority (reference: 20/HRA/210, IRAS: 282686) and local Trust Research and Development approval.

## Results

The total sample size was 4,378 (N=37,870), representing a response rate of 12%.

### Demographic composition of the sample in comparison to population

The demographic characteristics of the sample are described in Table 1. In all data presented here, frequencies are unweighted, and proportions are weighted to adjust for Trust size and non-response (Trust-level data provided by Trust HR: see Supplementary Table 1 for full weighted and unweighted proportions). There were some statistically significant differences between those who completed only the short survey (n=2,212), and those who completed both the long and short survey (n=2,166), with men, those identifying as being from racial and ethnic minority groups, those born outside the EU (including UK), and doctors, other clinical staff, and non-clinical staff being less likely than nurses to complete both surveys (see Supplementary Table 2 for full details).

**Table 1.** Demographic characteristics of the sample (n=4,378) ^a^.

| Characteristic |  | n (weighted %) |
| --- | --- | --- |
| Age (years) |  | Mean age: 41 (SD 12) |
|  | ≤30 | 985 (24.2) |
|  | 31-40 | 1,138 (30.7) |
|  | 41-50 | 979 (23.5) |
|  | 51-60 | 861 (20.8) |
|  | ≥61 | 171 (4.1) |
| Sex |  |  |
|  | Female | 3,482 (74.8) |
|  | Male | 833 (24.6) |
|  | Other | 6 (0.2) |
|  | Prefer not to say | 18 (0.5) |
| Relationship status |  |  |
|  | Married/civil partnership | 1,827 (42.2) |
|  | Co-habiting/in a relationship | 1,129 (23.2) |
|  | Divorced/separated/widowed | 251 (6.3) |
|  | Single | 1,114 (28.3) |
| Ethnicity |  |  |
|  | White | 3,215 (53.2) |
|  | Black/African/Caribbean/Black British | 373 (20.0) |
|  | Asian/Asian British | 477 (17.4) |
|  | Mixed/Multiple racial and ethnic groups | 173 (3.9) |
|  | Other racial and ethnic minority groups<sup>b</sup> | 90 (4.5) |
| Country of birth |  |  |
|  | UK | 2,974 (61.1) |
|  | EU (not UK) | 525 (9.8) |
|  | Other | 812 (29.2) |
| Length of time living in the UK (for those born outside the UK) |  |  |
|  | <1-2 years | 106 (7.8) |
|  | 3-5 years | 163 (10.9) |
|  | 6-10 years | 210 (14.5) |
|  | 11-20 years | 404 (32.5) |
|  | 21-29 years | 223 (17.1) |
|  | ≥30 years | 220 (16.5) |
|  | Prefer not to say | 7 (0.7) |
| Main role<sup>c</sup> |  |  |
|  | Doctor | 557 (12.8) |
|  | Nurse | 1,107 (26.7) |
|  | Other clinical | 1,306 (28.3) |
|  | Non-clinical | 1,356 (32.3) |

<sup>a</sup> Frequencies are unweighted, proportions are weighted. Numbers may not add up due to missing data.
<sup>b</sup> 'Other racial and ethnic minority groups' includes the options 'Arab' and 'Any other ethnic background'.
<sup>c</sup> In the 'Main role' variable, 'Other clinical' includes all other clinical roles, e.g. physiotherapist, psychologist, dietician, radiographer etc. 'Non-clinical' includes all other roles, e.g. administrative, domestic services, IT support, finance etc.

### Mental health measures

Table 2 shows the prevalence of probable CMDs, measured by the GHQ, with 58.9% (95%CI 58.1, 60.8) of participants meeting the cut-off score. In terms of secondary outcomes, 23.2% (95%CI 21.3, 25.8) met the cut-off score for probable anxiety (GAD-7), 27.3% (95%CI 25.3, 29.4) for probable depression (PHQ-9), 10.5% (95%CI 9.2, 11.9) for probable alcohol misuse (AUDIT), 30.2% (95%CI 28.1, 32.5) for probable PTSD (PCL-6), and the sample had a mean score of 15.5 (95%CI 15.1, 16.0) on the moral injury scale (MIES). A full breakdown of each measure by all demographics is given in Supplementary Table 3.

**Table 2.** Prevalence of adverse mental health outcomes^a^.

| Characteristic | n | n (% meeting cut-off score) [95%CI] | Mean (95%CI) |
| --- | --- | --- | --- |
| Probable common mental disorders (CMDs)<br>(GHQ: Cut-off $\geq 4$, scale range: 0-12) | 3,967 | 2,338 (58.9)<br>[58.1, 60.8] | 4.9 (4.7, 5.0) |
| Probable anxiety<br>(GAD-7: Cut-off $\geq 10$, scale range: 0-28) | 2,471 | 558 (23.2)<br>[21.3, 25.3] | 6.5 (6.3, 6.8) |
| Probable depression<br>(PHQ-9: Cut-off $\geq 10$, scale range 0-36) | 2,466 | 665 (27.3)<br>[25.3, 29.4] | 7.1 (6.8, 7.3) |
| Alcohol misuse<br>(AUDIT: Cut-off $\geq 8$, scale range 0-45) | 2,308 | 278 (10.5)<br>[9.2, 11.9] | 3.5 (3.3, 3.7) |
| Probable PTSD<br>(PCL-6: Cut-off $\geq 14$, scale range 6-30) | 2,447 | 715 (30.2)<br>[28.1, 32.5] | 11.9 (11.7, 12.2) |
| Moral injury<br>(MIES: No cut-off, scale range 9-54) | 2,399 | (no cut-off) | 15.5 (15.1, 16.0) |

<sup>a</sup> Frequencies are unweighted, proportions and means are weighted.
GHQ was measured in the short survey, while the other measures were included in the optional longer survey, hence the difference in numbers completing each.

In the past two months, 8.5% (95%CI 7.3, 9.8) of participants had considered taking their own life, while 2.0% (95%CI 1.4, 2.7) had attempted suicide, and 3.0% (95%CI 2.3, 3.9) had harmed themselves (see Table 3 for full details).

**Table 3.** Prevalence of suicidal thoughts (CIS-R) ^a^.

|  | n | Yes, in the past 2 months<br>n (%) [95%CI] | Yes, but not in the past 2 months<br>n (%) [95%CI] | No<br>n (%) [95%CI] |
| --- | --- | --- | --- | --- |
| Have you ever thought of taking your life, even if you would not really do it? | 2,487 | 227 (8.5)<br>[7.3, 9.8] | 526 (19.5)<br>[17.8, 21.3] | 1,734 (72.0)<br>[70.0, 74.0] |
| Have you ever made an attempt to take your life, by taking an overdose of tablets or in some other way? | 2,487 | 47 (2.0)<br>[1.4, 2.7] | 136 (5.1)<br>[4.2, 6.2] | 2,304 (92.9)<br>[91.7, 94.0] |
| Have you ever harmed yourself in any way but not with the intention of killing yourself? | 2,488 | 75 (3.0)<br>[2.3, 3.9] | 282 (9.6)<br>[8.4, 10.9] | 2,131 (87.4)<br>[85.9, 88.8] |

<sup>a</sup> Frequencies are unweighted, proportions are weighted.

### Logistic regression models

The multivariable logistic regression models showed that the likelihood of meeting the GHQ cut-off score, indicating probable CMDs, was 0.98 (95%CI 0.97, 0.99) times lower for each increased year of age, adjusting for all other covariates. Males were 0.7 (95%CI 0.5, 0.9) times less likely to have probable CMDs than females. Nurses were 2.2 (95%CI 1.5, 3.1) more likely to have probable CMDs than doctors, with other clinical and non-clinical staff also more likely to have probable CMDs than doctors. Those reporting high exposure to potentially morally injurious events were 2.6 (95%CI 2.0, 3.4) times more likely to report symptoms of probable CMDs than those reporting low exposure (Table 4).

**Table 4.** Weighted multivariable binary logistic regression models of factors associated with adverse mental health outcomes.

|  | Probable common<br>mental disorders<br>OR (95%CI)<br>n=2,245 | Probable anxiety<br>OR (95%CI)<br>n=2,273 | Probable depression<br>OR (95%CI)<br>n=2,278 | Probable alcohol misuse<br>OR (95%CI)<br>n=2,112 | Probable PTSD<br>OR (95%CI)<br>n=2,285 |
| --- | --- | --- | --- | --- | --- |
| Age | **0.98 (0.97, 0.99) \*\*\*** | **0.97 (0.96, 0.98) \*\*\*** | **0.97 (0.96, 0.98) \*\*\*** | 0.99 (0.98, 1.00) | **0.98 (0.97, 0.99) \*\*\*** |
| Sex (reference: female) |  |  |  |  |  |
| Male | **0.7 (0.5, 0.9) \*\*** | **0.7 (0.5, 0.9) \*\*** | 0.8 (0.6, 1.1) | 1.4 (1.0, 2.0) | **0.6 (0.5, 0.9) \*\*** |
| Other | 0.4 (0.1, 2.7) | 1.2 (0.3, 6.3) | 1.0 (0.2, 4.9) | - | 0.3 (0.0, 2.5) |
| Prefer not to say | 0.3 (0.0, 3.7) | - | 0.28 (0.03, 2.62) | - | - |
| Ethnicity (reference: White) |  |  |  |  |  |
| Black | 1.0 (0.7, 1.4) | 1.0 (0.7, 1.6) | **0.6 (0.4, 0.9) \*** | **0.3 (0.1, 0.6) \*\*** | 1.1 (0.7, 1.6) |
| Asian | 0.8 (0.6, 1.0) | 0.9 (0.6, 1.3) | 0.9 (0.7, 1.3) | **0.3 (0.1, 0.6) \*\*\*** | 0.9 (0.7, 1.3) |
| Mixed racial and<br>ethnic minority groups | 1.1 (0.7, 1.8) | 1.1 (0.7, 1.9) | 1.1 (0.6, 1.8) | 0.6 (0.3, 1.2) | 1.0 (0.6, 1.7) |
| Other racial and<br>ethnic minority groups<sup>a</sup> | 1.3 (0.6, 3.3) | 0.5 (0.2, 1.4) | 1.3 (0.5, 3.5) | **0.3 (0.1, 0.6) \*\*** | 0.7 (0.3, 2.1) |
| Role (reference: Doctor) |  |  |  |  |  |
| Nurse | **2.2 (1.5, 3.1) \*\*\*** | **1.9 (1.2, 3.1) \*\*\*** | **2.3 (1.5, 3.5) \*\*\*** | 1.0 (0.6, 1.8) | **1.9 (1.3, 2.9) \*\*\*** |
| Other clinical | **1.5 (1.1, 2.2) \*\*** | 1.2 (0.7, 1.9) | **1.6 (1.0, 2.5) \*** | 0.7 (0.4, 1.3) | 1.5 (0.9, 2.3) |
| Non-clinical | **1.5 (1.1, 2.2) \*\*** | **1.8 (1.2, 2.9) \*\*** | **2.5 (1.6, 3.8) \*\*\*** | 1.4 (0.8, 2.3) | **1.8 (1.2, 2.7) \*\*** |
| Moral injury (MIES) score<br>(reference: Low) |  |  |  |  |  |
| Medium | **1.3 (1.0, 1.7) \*** | **1.6 (1.1, 2.1) \*\*** | **1.4 (1.0, 1.8) \*** | 1.4 (0.9, 2.0) | **1.8 (1.4, 2.4) \*\*\*** |
| High | **2.6 (2.0, 3.4) \*\*\*** | **3.0 (2.2, 4.0) \*\*\*** | **2.9 (2.2, 3.9) \*\*\*** | **1.7 (1.1, 2.5) \*\*** | **4.2 (3.2, 5.6) \*\*\*** |

Differences within categories are statistically significant at: \*p<0.05, \*\*p<0.01, \*\*\*p<0.001.
Odds ratios shown are adjusted for all other covariates in each model.
a 'Other racial and ethnic minority groups' includes the options 'Arab' and 'Any other ethnic background'.

These patterns were similar when exploring our secondary outcomes of interest, with older participants and men being less likely to have probable anxiety and PTSD than younger participants and women. Black HCWs were less likely to have probable depression compared to their white peers, while Black, Asian, and other racial and ethnic minority groups were less likely to engage in alcohol misuse than white HCWs. Doctors were less likely to report symptoms of probable anxiety, depression and PTSD compared to all other groups of HCWs. Those with higher exposure to moral injury were more likely to have probable depression, alcohol misuse, and PTSD symptoms. Full details are shown in Table 4.

## Discussion

### Statement of principal findings

Our study found a high prevalence of adverse mental health outcomes amongst a sample of London-based HCWs during the first wave of the COVID-19 pandemic. Of particular note, we found concerning numbers of HCWs who reported symptoms of PTSD, and that higher exposure to moral injury was associated with worse outcomes. We identified groups at higher risk of experiencing poor mental health included women, younger staff, and nurses.

### Strengths and limitations

The NHS CHECK multi-site study provides findings from three different NHS Trusts, including acute and mental health services. The sites are located in London, which had a high regional case burden of COVID-19 during the first wave when recruitment commenced. This study represents a large sample size (n=4,378) relative to existing literature, with similar demographic characteristics to the overall NHS workforce in terms of age and sex [17], while London Trusts typically have a higher proportion of staff from racial and ethnic minority groups than other areas of the country, which is represented in our weighted data. We had detailed population-level HR information and were able to estimate response rates and the extent of non-response bias. These are notable methodological strengths compared with existing literature which have largely not reported response rates or assessed for related bias.[19] This study was uniquely inclusive in having gathered data from all staff employed by the NHS Trusts, including non-clinical, ancillary, lower-paid, and temporary staff. To our knowledge, this is also the only study that considers moral injury in the context of COVID-19 in HCWs.

However, there are some limitations. Firstly, despite substantial recruitment efforts as outlined above, the response rate was 12% and therefore it is inevitable that findings are open to selection bias, with those for whom the survey had greatest salience (i.e. those who were distressed) probably being most likely to participate. We hypothesise that the participation levels may be reflective of HCW burden related to COVID-19, such as work pressures and research/survey fatigue at this time, as well as longer term decline in HCW participation rates in surveys as observed over recent decades.[20] We found evidence that participants from some demographic and occupational groups, notably those from racial and ethnic minority groups and men, were less likely to participate. Although we took steps to weight the data for such non-participation, differential participation may lead to inaccuracies in prevalence estimates and hampers generalisability of the results. These concerns are compounded by differential response between our short and long surveys, however due to the time limitations on HCWs it was necessary to provide the option to complete only the short survey.

Secondly, it has also been shown that occupational specific surveys of any given group give rise to higher prevalence of mental health distress compared to population studies that contain sufficient numbers of the same occupational group for analysis, suggesting a possible contextual or framing effect.[21] Thirdly, given that all three sites were based in London, we have not captured associated national geographical variations.. Finally, the data was at this stage cross-sectional, meaning we were unable to look at whether symptoms persist, although as noted above, this is preliminary data, and 15 additional sites and longitudinal data will follow. Unfortunately, as this study began in response to the COVID-19 pandemic, we do not have pre-COVID baseline data with which to compare our results to, which is a significant limitation.

### Comparison with literature

#### International HCWs

Direct comparison of our findings with other COVID-19 HCW studies is challenging owing to varied methodological quality, and heterogeneity of healthcare systems in different countries, which is reflected in the wide range of results published to-date. A systematic review of 54,707 HCWs from 59 HCW specific international studies during COVID-19 found median anxiety levels of 24% (range 9- 90%), depression levels of 21% (range 5-51%) and general psychological distress of 37% (range 7- 97%), although measures and cut-off scores used in the included studies were heterogenous.[22] Compared to the reported median, our results show higher prevalence of poor psychological health, higher levels of depression, and similar levels of anxiety.

#### UK HCWs

In comparison to other UK based HCW studies, our results show lower levels of anxiety and depression, though higher levels of PTSD. A cross-sectional study (n=1194) using convenience sampling found 47.3% anxiety (using a lower cut-off than us of ≥8 on the GAD-7), 46.9% depression (using the same cut-off as us of ≥10), and 22.5% PTSD (using a different measure: ITQ,[23]).[24] A different cross-sectional study (n=2773) found levels (using the same cut-off scores of ≥10 for the GAD-7 and PHQ9, though a different PTSD measure; IES-R,[25]), of 33.1% for probable anxiety, 28.1% for probable depression, and 14.6% for PTSD symptoms.[26] Elevated prevalence of anxiety and depression compared to our study may be attributable to differing sample demographic factors, particularly sex, which was over-represented in these studies.

#### Previous epidemics

With the notable exception of PTSD, our results are comparable to findings from previous epidemics. A rapid systematic review and meta-analysis of the impact of viral epidemic outbreaks on the mental health of HCWs that included 117 international studies found a pooled prevalence of anxiety (30%), depression (24%), and PTSD (13%).[27] The level of probable PTSD in our study (30.2%) was more than double this level, however Serrano-Ripoll et al provide no information about the PTSD measures used in included studies, nor the cut-off scores used, making direct comparisons challenging. We used the recommended cut-off score of ≥14 on the PCL-6.[13]

#### General population

Contrasting our results with contemporary non-HCW specific population level studies conducted during the COVID-19 pandemic, which use the same measure (GHQ) and cut-off score (4), we report higher prevalence of poor general psychological health (58.9%) in HCWs. The Understanding Society (n=17,452)[6] and the UK Household Longitudinal Survey (n=12,074),[4] found prevalence of poor psychological health to be 27.3% and 37.1% respectively. Prevalence of anxiety and depression symptoms were similar to the “COVID-19 Social Study” (n=53,328),[5] and “Mental health in the UK and COVID-19 “study (n=3,097),[28] which found 24.4% and 26.0% participants respectively met the threshold for anxiety, and 31.4% and 31.6% respectively for depression (using the same measures and cut-off scores as us). However the demographic profile of these samples (where given) were different to ours, e.g. with more men (46.4%) and a higher average age (51 years),[5] and more white participants (93.6%,[5]; 95%,[6]).

#### Associated factors

Notably, some results from population studies large enough to include subsamples of HCWs did not find that HCWs had significantly different mental health to the general population and/or noted that any differences found might be explained by demographic differences between HCWs and the general population.[6,29] Others show an increase for keyworkers (samples which included HCWs) of anxiety [28,30] and depression.[30] Similarly, systematic reviews and meta-analyses that compared results from HCW and general population studies found varied results, with some suggesting elevated risk amongst HCWs,[31,32] whilst others did not find any statistically significant difference in depression, anxiety, PTSD and psychological distress related to HCW status.[33] As such, whilst there is good evidence of increased stress, depression, and anxiety across the general population and HCWs, robust evidence is still needed to determine whether the risk of mental-ill health is comparatively greater in HCWs.

The odds of having CMDs were greater amongst women, those of a younger age, and in specific occupational roles including nurses, other clinical and non-clinical staff (relative to doctors). Younger age was also associated with increased levels of anxiety, depression and PTSD. Female sex was associated with increased anxiety and PTSD. Nurses and non-clinical staff were disproportionately impacted by anxiety, depression and PTSD. These associations are in-keeping with literature from previous epidemics [27] and COVID-19,[22,34] whereby female sex, younger age, and specific occupational roles (e.g. nursing) are associated with increased adverse outcomes. Our findings contribute to this strong and evolving weight of evidence.

#### Self-harm and suicidality

Our study reported 8.5% of participants had considered taking their own life in the two previous months, while 2.0% had attempted suicide and 3.0% had harmed themselves in the same time period. A contemporary general population study in the UK, COVID-19 Social Study (n=44,775), reported data between March and April 2020 that found 18% experienced thoughts of suicide or self-harm (not directly comparable to our data) and 5% reported self-harm behaviour (higher than our rate).[35] A systematic review and meta-analysis (n=25 studies) of doctors conducted prior to the pandemic (1970s to 2018) found 17% suicidal ideation and 1% lifetime suicide attempts,[37] which is lower than UK population data, at 20.6% suicidal ideation, and 6.7% lifetime suicide attempts.[36] Overall, while any level of suicide ideation is concerning, our results do not provide evidence for elevated suicidal ideation, or acts of self-harm, compared to the general population, however it is possible that suicide attempts may be elevated. Interpretation of these findings must be cautious due to a lack of robust comparative data, and contextualised with current research findings from high income countries that have found no population-wide rise in suicide rates due to COVID-19 to date.[37]

#### Moral injury

HCWs with high exposure to moral injury had elevated odds of probable CMDs, depression, alcohol misuse, and PTSD. This is consistent with concerns during the pandemic [38] and previous occupational moral injury literature. A systematic review and meta-analysis, unrelated to COVID-19, that included 13 studies representing 6,373 participants found that exposure to potentially morally injurious experiences increased risk of PTSD, depression, and suicidality.[39] Although most studies included in the systematic review were from military samples, our study adds to the evidence that there may be an association between exposure to moral injury and mental disorders in HCW populations.

### Unanswered questions and future work

The wider NHS CHECK study, from which the data presented here were taken, is collecting data at baseline, with consent from participants to contact them for follow-up data at six-monthly intervals thereafter, dependent on how the pandemic evolves over time. We have expanded our study population to ensure a broader, more representative geographical spread, and our study now includes 18 sites across England and Wales. In particular, the elevated rates of PTSD identified warrant further investigation in the ongoing study and larger data-set analysis. As data from more sites become available, we will also explore the impact of organisational and social factors on the experiences of staff and, mental health outcomes (e.g. working practices, perceived support from colleagues and managers, COVID-19 impacts and experiences, and staff support programmes available and used).

Further, although we have used validated mental health questionnaires, questionnaire-based surveys of mental health are case-finding, rather than diagnostic tools, which favour sensitivity over specificity. Occupational self-report measures commonly overestimate rates of “caseness”, i.e. individuals who are likely to meet true diagnostic criteria and might thus require specific interventions.[21] Collecting information about distress at a time of great stress means that high proportions of participants reporting difficulties might be expected, however this is not necessarily indicative of being mentally ill. In particular, we need to know whether these symptoms persist and whether they are associated with a reduction in ability to function at work and home, and other meaningful outcomes, for example, help seeking, reduced desire to remain in their post. Hence we will be extending the study in a two stage psychiatric epidemiology design,[40] using standard psychiatric diagnostic interviews administered by phone with a sub-sample of participants, in order to distinguish distress from disorder.

We anticipate finishing baseline data collection in December 2020, with follow-up data collection starting at six-months post-baseline for each individual participant. We will continue to increase response rates through extra engagement tools, including: new patient and public involvement (PPI) initiatives, further work with trade union representatives, offering incentives to participants, and ensuring paper surveys are available in addition to the online version. We will also focus on engaging with racial and ethnic minority groups to address the relatively reduced participation rate and completion of the longer component of the survey by forming partnerships with those experienced in such engagements. Inclusion of an ethnicity-focused module at eight months will help capture inequalities in mental health and occupational outcomes.

### Implications

HCWs are experiencing a high burden of adverse mental health outcomes during the COVID-19 pandemic and some demographic and occupational groups are more vulnerable than others. The mental health offering to HCWs must therefore consider the overlap of these factors. For example, our findings indicate that young female nurses may be more vulnerable to adverse mental health outcomes and therefore may benefit from targeted support.

Despite the high levels reported by our study, and others, there remains uncertainty about whether HCWs have worse COVID-19 related mental health outcomes compared to the general population. We urge additional research that emphasises quality and methodological rigor, namely with use of standardised diagnostic interviews with a defined sampling frame. In doing so, the mental health offer will be better able to avoid pathologizing ordinary distress responses, identify those most at risk, and provide necessary support.

## Conclusion

Our preliminary data indicate that continued monitoring of HCWs is vital in order to identify those groups most at risk of adverse mental health outcomes. Longitudinal data and diagnostic interviews will be needed to pick apart mental disorders from normal distress responses, and ensure appropriate support is provided to those most in need. Our findings suggest that it is not only clinical staff who may need support, but the full range of HCWs contributing to the pandemic effort.

## Data Availability

All data requests should be submitted to the corresponding author for consideration. Access to available anonymised data may be granted following review.

## Data Availability

We used data from the Avon Longitudinal Study of Parents and Children (ALSPAC), an ongoing population-based study that contains a wide range of phenotypic and environmental measures, genetic information and linkage to health and administrative records.

http://www.bristol.ac.uk/alspac/researchers/our-data/

http://www.bris.ac.uk/alspac

## Contributor and guarantor information

DLa and SG wrote the first draft of the manuscript. DLa and EC carried out data analysis. RB managed the study. SAMS, SW, NG, MH, ReR, and RoR are co-chief investigators. SC, AD, MD, SD, SG, SLH, CWJ, DLa, DLe, IMa, SM, IMc, AMR, MP, and CP are co-investigators. PA, VF, HG, JMB, RH, and CS are site leads. DS designed the data collection tool. All authors contributed edits to the manuscript.

The corresponding author attests that all listed authors meet authorship criteria and that no others meeting the criteria have been omitted.

## Copyright

The Corresponding Author has the right to grant on behalf of all authors and does grant on behalf of all authors, a worldwide licence to the Publishers and its licensees in perpetuity, in all forms, formats and media (whether known now or created in the future), to i) publish, reproduce, distribute, display and store the Contribution, ii) translate the Contribution into other languages, create adaptations, reprints, include within collections and create summaries, extracts and/or, abstracts of the Contribution, iii) create any other derivative work(s) based on the Contribution, iv) to exploit all subsidiary rights in the Contribution, v) the inclusion of electronic links from the Contribution to third party material where-ever it may be located; and, vi) licence any third party to do any or all of the above.

## Competing interests declaration

MH, RR, and SW are senior NIHR Investigators. This paper represents independent research part-funded by the NIHR Maudsley Biomedical Research Centre Trust and King’s College London (MH, SW, SAMS). The views expressed are those of the authors and not necessarily those of the NHS, the NIHR, or the Department of Health and Social Care.

### From ICMJE forms

Prof Raine reports grants from DHSC/UKRI/ESRC COVID-19 Rapid Response Call, grants from Rosetrees Trust, grants from King’s Together rapid response call, grants from UCL (Wellcome Trust) rapid response call, during the conduct of the study; & grants from NIHR outside the submitted work.

Dr. Hotopf reports grants from DHSC/UKRI/ESRC COVID-19 Rapid Response Call, grants from Rosetrees Trust, grants from King’s Together rapid response call, grants from UCL Partners rapid response call, during the conduct of the study; grants from Innovative Medicines Initiative and EFPIA, RADAR-CNS consortium, grants from MRC, grants from NIHR, outside the submitted work.

Professor Hatch reports grants from NIHR, grants from Wellcome Trust, grants from ESRC, grants from Guy’s and St. Thomas’ Charity, grants from MRC, grants from UKRI, outside the submitted work; and I am a member of the following advisory groups: The Health Foundation - COVID-19 Research Programme Panel, NHS England and NHS Improvement - Patient and Carers Race Equalities Framework [PCREF] Steering Group, NHS England and NHS Improvement - Advancing Mental Health Equalities Taskforce, Health Education England - Mental Health Workforce Equalities Subgroup, Maudsley Learning - Maudsley Learning Advisory Board, South London and Maudsley NHS Foundation Trust (SLaM) - Independent Advisory Groups, the SLaM Partnership Group, Lambeth Public Health - Serious Youth Violence Public Health Task and Finish Group, NHS England - Workforce Race Equality Standard Advisory Group, Thrive London - Thrive London Advisory Board, Black Thrive - Black Thrive Advisory Board. Commissions: Welsh Government’s Race Equality Plan; contribution to the evidence review for Health and Social Care and Employment and Income policy areas.

Dr. Stevelink reports grants from UKRI/ESRC/DHSC, grants from UCL, grants from UKRI/MRC/DHSC, grants from Rosetrees Trust, grants from King’s Together Fund, during the conduct of the study.

Prof. Greenberg reports a potential COI with NHSEI, during the conduct of the study; and I am the managing director of March on Stress Ltd which has provided training for a number of NHS organisations although I am not clear if the company has delivered training to any of the participating trusts or not as I do not get directly involved in commissioning specific pieces of work.

Other authors report no competing interests.

## Funding

NHS CHECK has received funding from the following organisations and charities for the period of data collection reported in this manuscript: National Institute for Health Research Maudsley Biomedical Research Centre, King’s College London; Rosetrees Trust; and the National Institute for Health Research Health Protection Research Unit in Emergency Preparedness and Response at King’s College London. The funders had no role in the design, analysis, interpretation or decision to submit this paper. The joint first authors had full access to the data in the study and had final responsibility, with endorsement from the joint last authors (SW and SAMS), for the decision to submit the paper for publication. The views expressed are the views of the authors and do not necessarily represent the views of their organisations or funding sources.

RoR and DLa are funded by the National Institute for Health Research (NIHR) Applied Health Research North Thames (NIHR ARC North Thames). This funder had no role in study design, data collection, data analysis, data interpretation, or writing of the report. The views expressed in this article are those of the authors and not necessarily those of the NHS, the NIHR, or the Department of Health and Social Care.

## Acknowledgements

We are especially grateful to all the participants who took part in the study. We would also like to extend our thanks to Haifa Issa, Dr Howard Burdett, Melanie Chesnokov, the medical students who helped to recruit participants at the research sites and all NHS staff who promoted NHS CHECK. We would also like to thank the TIDES study team for their support and collaboration.

RoR and DLa are support by the National Institute for Health Research (NIHR) Applied Health Research North Thames (NIHR ARC North Thames). This funder had no role in study design, data collection, data analysis, data interpretation, or writing of the report. The views expressed in this article are those of the authors and not necessarily those of the NHS, the NIHR, or the Department of Health and Social Care.

