## Supplemental tables 1, 2, 3 for "The psychosocial impact of the COVID-19 pandemic on 4,378 UK healthcare workers and ancillary staff: initial baseline data from a cohort study collected during the first wave of the pandemic"

### Supplementary tables

**Supplementary table 1 Weighted and unweighted socio-demographics (n=4,378)**

| **Variable** | | **n** | **Unweighted %** | **Weighted %** |
| --- | --- | --- | --- | --- |
| Age (years) | |  |  |  |
|  | ≤30 | 985 | 23.8 | 24.2 |
|  | 31-40 | 1,138 | 27.5 | 30.7 |
|  | 41-50 | 979 | 23.7 | 23.5 |
|  | 51-60 | 861 | 20.8 | 20.8 |
|  | ≥61 | 171 | 4.1 | 4.1 |
| Sex | |  |  |  |
|  | Female | 3,485 | 80.3 | 74.8 |
|  | Male | 833 | 19.2 | 24.6 |
|  | Other | 6 | 0.1 | 0.2 |
|  | Prefer not to say | 18 | 0.4 | 0.5 |
| Relationship status | |  |  |  |
|  | Married/Civil partnership | 1,827 | 42.3 | 42.2 |
|  | Co-habiting/in a relationship | 1,129 | 26.1 | 23.2 |
|  | Divorced/separated/widowed | 251 | 5.8 | 6.3 |
|  | Single | 1,114 | 25.8 | 28.3 |
| Ethnicity | |  |  |  |
|  | White | 3,263 | 74.5 | 53.5 |
|  | Black/African/Caribbean/Black British | 373 | 8.5 | 20.8 |
|  | Asian/Asian British | 482 | 11.0 | 17.4 |
|  | Mixed/Multiple racial and ethnic minority groups | 173 | 4.0 | 3.8 |
|  | Other racial and ethnic minority groups^a^ | 90 | 2.1 | 4.5 |
| Country of birth | |  |  |  |
|  | UK | 2,974 | 68.9 | 61.0 |
|  | EU not UK) | 525 | 12.2 | 9.8 |
|  | Other | 815 | 18.9 | 29.2 |
| Length of time living in the UK | |  |  |  |
|  | <1-2 years | 106 | 8.0 | 7.8 |
|  | 3-5 years | 163 | 12.2 | 10.9 |
|  | 6-10 years | 210 | 15.8 | 14.5 |
|  | 11-20 years | 404 | 30.3 | 32.5 |
|  | 21-29 years | 223 | 16.7 | 17.1 |
|  | >30 years | 220 | 16.5 | 16.5 |
|  | Prefer not to say | 7 | 0.5 | 0.7 |
| Main role | |  |  |  |
|  | Doctor | 557 | 12.9 | 12.8 |
|  | Nurse | 1,108 | 25.6 | 26.7 |
|  | Other clinical | 1,306 | 30.2 | 28.3 |
|  | Non-clinical | 1,358 | 31.4 | 32.3 |

a ‘Other racial and ethnic minority groups’ includes the options ‘Arab’ and ‘Any other ethnic background’.

**Supplementary table 2 Socio-demographics of short-survey-only and short-and-long-survey participants**

| **Variable** | | **Short-survey only**  **n (%)^a^** | **Short- and long-survey**  **n (%)^a^** | **Significant differences**  **p=** |
| --- | --- | --- | --- | --- |
| Age (years) | |  |  |  |
|  | ≤30 | 464 (23.4) | 521 (25.1) | 0.31 |
|  | 31-40 | 574 (31.0) | 564 (30.4) |  |
|  | 41-50 | 522 (24.9) | 457 (22.0) |  |
|  | 51-60 | 419 (16.7) | 442 (18.5) |  |
|  | ≥61 | 86 (4.0) | 85 (4.0) |  |
| Sex | |  |  |  |
|  | Female | 1,691 (72.1) | 1,791 (74.8) | 0.001 |
|  | Male | 473 (27.5) | 360 (24.6) |  |
|  | Other | 1 (<1) | 5 (<1) |  |
|  | Prefer not to say | 10 (<1) | 8 (<1) |  |
| Relationship status | |  |  |  |
|  | Married/Civil partnership | 930 (42.3) | 897 (42.1) | 0.24 |
|  | Co-habiting/in a relationship | 538 (21.9) | 591 (24.7) |  |
|  | Divorced/separated/widowed | 128 (6.8) | 123 (5.9) |  |
|  | Single | 566 (29.1) | 546 (27.4) |  |
| Ethnicity | |  |  |  |
|  | White | 1,530 (48.4) | 1,685 (59.4) | <0.0001 |
|  | Black/African/Caribbean/Black British | 219 (22.7) | 154 (19.0) |  |
|  | Asian/Asian British | 283 (20.0) | 194 (14.6) |  |
|  | Mixed/Multiple racial and ethnic minority groups | 83 (3.5) | 90 (4.3) |  |
|  | Other racial and ethnic minority groups^b^ | 55 (5.3) | 35 (3.7) |  |
| Country of birth | |  |  |  |
|  | UK | 1,440 (58.2) | 1,534 (64.2) | 0.0007 |
|  | EU (not UK) | 267 (9.6) | 258 (10.0) |  |
|  | Other | 454 (32.2) | 358 (25.8) |  |
| Length of time living in the UK | |  |  |  |
|  | <1-2 years | 59 (8.1) | 47 (7.4) | 0.49 |
|  | 3-5 years | 85 (10.9) | 78 (11.0) |  |
|  | 6-10 years | 102 (12.9) | 108 (16.7) |  |
|  | 11-20 years | 230 (33.9) | 174 (30.7) |  |
|  | 21-29 years | 120 (17.1) | 103 (17.2) |  |
|  | >30 years | 117 (16.2) | 103 (16.8) |  |
|  | Prefer not to say | 5 (1.0) | 2 (<1) |  |
| Main role | |  |  |  |
|  | Doctor | 316 (13.9) | 241 (11.7) | 0.001 |
|  | Nurse | 479 (23.7) | 628 (29.9) |  |
|  | Other clinical | 691 (29.7) | 615 (26.8) |  |
|  | Non-clinical | 677 (32.8) | 679 (32.3) |  |

a Numbers are unweighted, proportions are weighted.
b ‘Other racial and ethnic minority groups’ includes the options ‘Arab’ and ‘Any other ethnic background’.

Supplementary table 3 Prevalence of mental health outcomes by socio-demographic factors ^a^

| **Characteristic** | | **Probable common mental disorders % (95%CI)** | **Probable anxiety % (95%CI)** | **Probable depression % (95%CI)** | **Probable alcohol misuse % (95%CI)** | **Probable PTSD % (95%CI)** | **Perceived moral injury Mean (95%CI)** |
| --- | --- | --- | --- | --- | --- | --- | --- |
| **Age years)** | | **n=3,785***** | **n=2,369***** | **n=2,368***** | n=2,219 | **n=2,351***** | **n=2,304 **** |
|  | ≤30 | 67.5 [63.8, 70.9] | 31.3 (27.0, 35.9) | 35.3 (30.9, 40.1) | 11.7 (9.1, 15.0) | 35.8 (31.4, 40.5) | 16.0 (15.2, 16.8) |
|  | 31-40 | 60.8 [57.2, 64.2] | 24.2 (20.6, 28.1) | 30.6 (26.6, 34.8) | 11.0 (8.7, 13.9) | 32.3 (28.3, 36.5) | 15.9 (15.0, 16.7) |
|  | 41-50 | 58.5 [54.4, 62.4] | 22.0 (18.0, 26.5) | 23.6 (19.6, 28.2) | 9.2 (7.0, 12.1) | 30.6 (25.9, 35.7) | 15.6 (14.7, 16.5) |
|  | 51-60 | 49.9 [45.6, 54.2] | 17.7 (13.9, 22.3) | 20.7 (16.7, 25.3) | 10.0 (7.3, 13.5) | 21.9 (17.8, 26.6) | 14.5 (13.7, 15.2) |
|  | ≥61 | 43.0 [33.1, 53.5] | 6.8 (3.4, 13.1) | 12.7 (7.4, 20.9) | 8.9 (4.4, 17.2) | 19.3 (11.2, 31.3) | 14.4 (12.5, 16.2) |
| **Sex** | | **n=3,965***** | **n=2,471*** | n=2,466 | =2,308 | **n=2,447**** | n=2,399 |
|  | Female | 62.2 (60.2, 64.3) | 24.8 (22.6, 27.1) | 28.8 (26.5, 31.2) | 9.5 (8.2, 11.0) | 32.4 (30.0, 35.0) | 15.3 (14.9, 15.7) |
|  | Male | 48.7 (44.5, 52.9) | 18.2 (14.4, 22.7) | 22.5 (18.4, 27.3) | 13.8 (10.7, 17.6) | 23.6 (19.2, 28.5) | 16.3 (15.3, 17.4) |
|  | Other | 58.6 (19.3, 89.3) | 32.3 (7.0, 75.0) | 32.3 (7.1, 75.0) | 0 | 15.9 (2.0, 63.5) | 19.2 (9.7, 28.8) |
|  | Prefer not to say | 67.0 (34.7, 88.6) | 23.9 (4.9, 65.7) | 30.2 (7.5, 70.0) | 0 | 9.8 (1.2, 50.4) | 14.1 (9.1, 19.1) |
| **Relationship status** | | n=3,953 | **n=2,463*** | **n=2,459***** | **n=2,301***** | **n=2,440***** | **n=2,392**** |
|  | Married/Civil partnership | 55.7 (52.8, 58.5) | 20.0 (17.2, 23.0) | 21.9 (19.1, 25.0) | 8.6 (7.0, 10.6) | 25.1 (22.1, 28.4) | 14.9 (14.3, 15.5) |
|  | Co-habiting/in a relationship | 61.3 (57.7, 64.8) | 26.5 (22.9, 30.5) | 30.4 (26.5, 34.5) | 14.2 (11.5, 17.6) | 34.8 (30.7, 39.2) | 15.6 (14.8, 16.3) |
|  | Divorced/separated/widowed | 59.8 (51.5, 67.6) | 21.4 (14.6, 30.1) | 23.3 (16.4, 32.0) | 3.6 (1.6, 7.7) | 39.7 (30.1, 50.1) | 16.3 (14.8, 17.8) |
|  | Single | 61.9 (58.2, 65.4) | 25.6 (21.6, 30.1) | 33.7 (29.3, 38.4) | 11.4 (8.8, 14.5) | 32.1 (27.8, 36.6) | 16.4 (15.5, 17.3) |
| **Ethnicity** | | n=3,958 | n=2,466 | n=2,461 | **n=2,303***** | n=2,442 | **n=2,394***** |
|  | White | 59.6 (57.6, 61.4) | 22.6 (20.7, 24.7) | 27.5 (25.4, 29.7) | 14.5 (12.8, 16.4) | 29.5 (27.3, 31.7) | 14.9 (14.5, 15.4) |
|  | Black/African/ Caribbean/Black British | 58.4 (52.8, 63.9) | 24.7 (18.5, 32.1) | 21.9 (16.0, 29.1) | 4.1 (1.9, 8.4) | 31.1 (24.3, 38.97) | 15.3 (14.1, 16.4) |
|  | Asian/Asian British | 55.9 (50.8, 60.8) | 24.2 (18.9, 30.5) | 30.6 (24.6, 37.2) | 4.9 (2.5, 9.3) | 32.1 (26.1, 38.8) | 18.0 (16.5, 19.5) |
|  | Mixed/Multiple ethnic groups | 65.8 (57.7, 73.0) | 26.4 (18.3, 36.5) | 31.3 (22.4, 41.7) | 8.7 (4.3, 16.9) | 33.2 (24.2, 43.5) | 15.5 (13.8, 17.1) |
|  | Other racial and ethnic minority groups | 59.6 (46.7, 71.3) | 17.0 (8.4, 31.5) | 33.3 (18.9, 51.7) | 0 | 25.8 (13.4, 43.9) | 16.3 (13.4, 19.2) |
| **Country of birth** | | n=3,943 | =2,456 | n=2,451 | **n=2,293***** | n=2, 432 | n=2,385 |
|  | UK | 58.7 (56.5, 60.9) | 22.7 (20.5, 25.0) | 26.0 (23.7, 28.4) | 13.0 (11.4, 14.9) | 29.1 (26.7, 31.7) | 15.2 (14.8, 15.7) |
|  | EU not UK) | 63.9 (59.1, 68.4) | 23.6 (18.8, 29.3) | 29.7 (24.4, 35.7) | 7.1 (4.4, 11.4) | 33.5 (27.7, 39.8) | 16.2 (15.0, 17.3) |
|  | Other | 58.0 (53.7, 62.2) | 24.5 (19.8, 29.8) | 29.5 (24.6, 35.0) | 4.8 (3.1, 7.3) | 32.0 (28.1, 32.5) | 16.1 (15.1, 17.2) |
| **Length of time living in the UK** | | n=1,176 | n=674 | n=672 | n=627 | n=666 | **n=646**** |
|  | <1-2 years | 69.4 (58.4, 78.6) | 29.1 (17.4, 44.4) | 40.3 (27.0, 55.2) | 7.0 (2.9, 16.3) | 34.8 (22.5, 49.6) | 18.4 (15.3, 21.5) |
|  | 3-5 years | 68.7 (59.8, 76.5) | 27.1 (18.6, 37.6) | 28.6 (19.8, 39.3) | 4.4 (1.8, 10.5) | 26.8 (18.4, 37.4) | 17.8 (15.2, 20.3) |
|  | 6-10 years | 57.6 (49.4, 65.4) | 25.6 (17.8, 35.4) | 29.7 (21.5, 39.5) | 8.3 (4.3, 15.3) | 29.7 (21.4, 39.7) | 16.0 (13.7, 18.3) |
|  | 11-20 years | 60.5 (54.3, 66.5) | 24.2 (17.5, 32.6) | 27.1 (20.4, 35.0) | 5.3 (2.8, 9.9) | 37.7 (29.8, 46.2) | 16.0 (14.4, 17.5) |
|  | 21-29 years | 55.1 (46.6, 63.3) | 20.2 (13.0, 30.1) | 32.8 (23.0, 44.4) | 5.7 (2.5, 12.5) | 26.2 (17.8, 36.9) | 15.6 (13.9, 17.2) |
|  | >30 years | 50.8 (42.4, 59.2) | 19.1 (11.6, 29.9) | 23.2 (14.5, 35.0) | 2.4 (0.8, 7.3) | 30.0 (20.3, 42.0) | 14.5 (13.1, 15.9) |
|  | Prefer not to say | 61.5 (20.0, 91.1) | 35.0 (4.6, 85.6) | 35.1 (4.7, 85.7) | 7.0 (2.9, 16.3) | 70.7 (17.9, 96.4) | 14.6 (10.0, 19.1) |
| **Main role** | | **n=3,964 ***** | **n=2,480 ***** | **n=2,465***** | n=2,307 | **n=2,446***** | **n=2,398**** |
|  | Doctor | 46.9 (41.8,52.0) | 13.6 (9.7, 18.9) | 15.4 (11.4, 20.4) | 10.2 (6.9, 14.8) | 18.3 (13.8, 23.8) | 14.4 (13.4, 15.4) |
|  | Nurse | 68.1 (64.6,71.3) | 30.2 (26.3, 34.5) | 33.1 (29.0, 37.4) | 10.2 (8.0, 12.8) | 36.5 (32.3, 40.9) | 17.4 (16.5, 18.3) |
|  | Other clinical | 59.5 (56.1,62.9) | 19.5 (16.3, 23.2) | 24.0 (20.5, 28.2) | 8.2 (6.3, 10.7) | 29.0 (25.2, 33.3) | 15.2 (14.4, 16.0) |
|  | Non-clinical | 55.6 (52.2,59.1) | 23.8 (20.4, 27.5) | 29.3 (25.6, 33.2) | 12.6 (10.2, 15.5) | 30.0 (28.1, 32.5) | 14.6 (14.0, 15.3) |

a Numbers are unweighted, proportions are weighted.

Differences within categories are statistically significant at: *p<0.05, **p<0.01, ***p<0.001
